# Clinico-microbiological profile and treatment outcomes in patients with isoniazid monoresistant tuberculosis in South India

**DOI:** 10.1101/2023.01.31.23285245

**Authors:** M Venkata Prasanna, R Muthuraj, R Vimal Raj, S Govindarajan, R Pajanivel

## Abstract

Isoniazid (INH) is an important first-line medication for the treatment of tuberculosis. The impact that tuberculosis drug resistance has on treatment outcomes is a topic that is receiving a lot of attention these days because of the rising incidence of INH-resistant cases. Study involves a single group of patients who have been diagnosed with Isoniazid monoresistant tuberculosis. Treatment history and demographic data of the patients were obtained after informed consent. The mutation patterns of isoniazid were observed after multiplex PCR and Line Probe Assay (LPA). A total of 101 patient (M,F) records at the IRL, Puducherry were analyzed. The predominant gene responsible for TB was KATG (67.3%). The KATG Mut1 was a prime mutation observed in the present study population (58.41%). Study showed positive association with males (74%), occupation as coolie (88%), diabetes as comorbidity (33%), pulmonary tuberculosis as the TB site (98.01%), history of previous ATT intake in 43 patients (42.6%), katG mutation (67.3%), katG Mut 1 was the prime mutation (58.4%).The cure rate was high in *INH high concentration resistance* patients which was statistically significant (p=0.0167). INH monoresistance mutations seen in 64.3% of the patients with katG, compared to inhA (34.65%). Similar to katG mutations, inhA mutations also have MUT1 as their most frequent gene pattern. There is a significant association between males, diabetes, smoking and alcohol addictions were associated with high risk of developing high dose INH monoresistance (katG). High prevalence of recurrent tuberculosis was seen in high dose INH monoresistance tuberculosis. Patients who are microbiologically confirmed pulmonary tuberculosis and diabetes with rifampicin sensitive status needs to be checked for LPA for isoniazid sensitivity status to prevent treatment failure and relapse. It is crucial to understand the gene pattern in each of these patients since these mutations are closely associated to high or low-degree resistance to INH

## INTRODUCTION

One of the leading causes of death in developing and underdeveloped Countries is Ttuberculosis (TB), which is caused by the bacteria *Mycobacterium tuberculosis* (MTB). According to the most recent statistics provided by the World Health Organization (WHO), there were 6.4 million newly diagnosed cases of Tuberculosis in the year 2021, a drop in incidence attributed to under reporting due to covid 19 pandemic [1] although TB most frequently affects the lungs, it is also capable of affecting other areas of the body. The incidence of TB is higher in India and claims the lives of over half a million people [2]. Today, India has the highest number of cases of MTR-TB in the world and is responsible for one-fourth of the worldwide burden of the disease and has much lower success rates [3]. There is evidence that tuberculosis is gradually decreasing, the worldwide emergence and spread of Multi Drug Resistance (MDR) have become a major obstacle [4]. The impact of Tuberculosis drug resistance and treatment outcomes has been a cause of concern in the past few years because of the rise in incidence of INH-resistant. however, new data indicates that more than half of patients with INH-monoresistant TB may require a treatment course lasting more than 6 months [5]. Additionally, numerous reports have mentioned significantly varied treatment plans for INH monoresistant TB [6]. Mutations in katG or InhA regulatory genes are major genes responsible for INH resistance. Catalase peroxidase, an enzyme that changes INH into its physiologically active form, is encoded by the katG gene. INH was ineffective for treating *Mycobacterium tuberculosis* with this mutation profile because katG mutations, particularly those at codon 315, provide high levels of INH resistance. The principal target of active INH, nicotin-amide adenine dinucleotide dependent enoyl-acyl carrier protein reductase, as well as ethionamide (ETH) and prothionamide, are all encoded by the InhA regulatory region (PTH) [8]. High doses of INH may be effective against *M. tuberculosis* with InhA mutations result in low-level resistance to the medication [9]. The critical concentrations of INH resistance were categorized as low and high dose with cut off values <0.2 3g/ml and >1 g/ml respectively. Low and high concentration INH resistance can be viewed of as two separate entities because different genetic changes are assumed to be the cause of each [10].

Line probe assay (LPA) was developed in response to the MDR-TB crisis in the world and to identify drug resistance in TB patients. Through the National tuberculosis elimination program, LPA is a quick method based on polymerase chain reaction (PCR) that is used to identify *Mycobacterium tuberculosis* complex as well as drug sensitivity to rifampicin (RPM), isoniazid (INH) and other first line drugs. LPA only examines sputum samples that have an AFB smear positive result [11]. Comparing MDR-TB treatment to non-MDR-TB treatment, unsatisfactory outcomes were more connected with MDR-TB treatment [12]. WHO has established a brief treatment regimen for MDR-TB patients in an effort to overcome some of these issues [13]. Numerous studies have been conducted in different nations to identify the factors that affect MDR-TB treatment outcomes in both the general population and particular target populations, including adults, children, with comorbidities and those who have co-infected with HIV. These investigations have revealed that there are significant regional differences in the predictors of treatment outcome. Body Mass Index (BMI) >18.5kg/m2, use of more than four effective medications, a negative baseline sputum smear, and experiencing a surgical resection are some of the variables that were discovered to be related with favourable treatment outcomes [14]. Other factors which included the use of linezolid [15] or fluoroquinolones, customized treatment,[16] getting any help from the TB program, having more understanding about the disease, and having more faith and support from nurses and Physicians [9]. Other factors were co-infection with HIV, a positive smear at the beginning of therapy, a history of TB treatment, smoking, pre-XDRTB, age >44 years, Ofloxacin resistance, male sex, low body weight at diagnosis (40 kg), poor treatment adherence, smear positive at the second month of treatment, the use of conventional medicine, and treatment interruptions longer than 14 days [17–19]. Contradictory results have been found for some of the above-mentioned criteria in trials with unsatisfactory treatment outcomes, though [20] furthermore, there are still gaps in the literature regarding the differences between patients with low- and high-concentration monoresistant TB in terms of baseline traits, therapeutic regimen, side events, and outcomes [9]. In the present investigation, it is proposed a) to study the clinical profile of patients registered for Drug-Resistant Tuberculosis treatment within Puducherry tuberculosis unit by review of treatment registry. (b). To study the Isoniazid drug resistance pattern and the pattern of resistance mutation in these patients by review of Intermediate Reference laboratory (IRL) registry. ©. To assess the treatment outcomes of the INH monoresistant cases as recorded in the DR-TB treatment registry. (d). To correlate the clinical and microbiological profiles with treatment outcomes among patients with Isoniazid monoresistant tuberculosis within the Puducherry tuberculosis unit.

## Materials and methods

### Study design & Population

It is a retrospective record-based study of 100 patients. Patients who had been enrolled in IRL and DRTB registry to have Isoniazid monoresistant pulmonary tuberculosis in Intermediate Reference laboratory, Puducherry. Study involves single group of patients who were diagnosed with Isoniazid monoresistant tuberculosis.

### Eligibility criteria

#### Inclusion criteria

All the patients who were registered in Intermediate reference laboratory and Drug resistant tuberculosis registry as Isoniazid monoresistant tuberculosis. Patients who had been registered to have mutations in katG gene and inhA gene.

#### Exclusion criteria

Patients who had been already diagnosed to have Poly Drug-Resistant Tuberculosis, Multi-Drug Resistant Tuberculosis, and Extensive Drug Resistant Tuberculosis. Patient records with incomplete information for the study

#### Sample Size

All Isoniazid monoresistant patients with above-mentioned inclusion criteria enrolled in Intermediate reference lab (IRL) from 2014-2020.

**Minimum sample size is calculated to be 97** using the below mentioned formula.

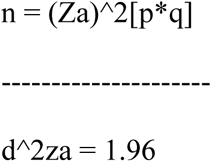

Estimated proportion (p)=0.5, q=1-p 1-0.5 =0.5

Estimated error (d)=10% (0.1)

**Sample size (n) =97**

### Study Tools

To assess phenotype and genotype characteristics of patients diagnosed with Isoniazid monoresistant tuberculosis, treatment regimens, treatment failure with the standard regimen and further how many were labelled from Isoniazid monoresistant tuberculosis to poly resistant tuberculosis, multidrug resistance and extensive drug resistant tuberculosis.

### Method of Data Collection

Patients diagnosed with Isoniazid mono-resistant tuberculosis were identified by the collection of data from Intermediate reference laboratory and drug resistant tuberculosis registry, Puducherry. Baseline characteristics/parameters to be included are as follows:

- Sex
- Age
- Comorbidities
- Smear positive Status
- Genes associated with the resistance
- Type of resistance – low and high dose Isoniazid resistance.
- History of treatment-new and previously treated. Previously treated case can be relapse (recurrent), failure or default (treatment after default).
- Radiographic Findings – Cavitary lesion, Bilateral lesion, Extensive lesion.

Treatment success and failure in each of the above Characteristics/Parameters. Data will be entered on Microsoft Excel sheets, checked for accuracy and completeness, and statistically analyzed to evaluate the significance of each risk factor in the development of Isoniazid monoresistant pulmonary tuberculosis.

### Data Processing and Statistical Analysis

The data was entered with an excel sheet. Data was exported to Medcalc version 19.2.6 [21] for further processing. All categorical variables were expressed as percentages and he continuous variables were expressed as mean ± standard deviation. The statistical significance of mean differences was compared in two groups using a independent t test and categorical variables were analysed using chi square test. All values were considered significant if the *p*-value was < 0.05.

### Ethical Consideration

This research was strictly fulfilling the ethical guidelines as outlined in the declaration of Helsinki, participants signed a consent form, and were assured that their participation was completely voluntary and could be terminated at any time without compromising their medical care. The study protocol was approved by the Mahatma Gandhi Medical College & Research Institute (MGMCRI) Review board.

## RESULTS

### Demographic and Clinical Characteristics of Patients

A total of 101 patients diagnosed with Isoniazid mono-resistant tuberculosis were identified by the collection of data from the Intermediate reference laboratory and drug resistance tuberculosis registry, Puducherry from 2014-2020 was included in the study. The baseline demographics and clinical characteristics of these patients are listed in Table 1.

**Table 1:**
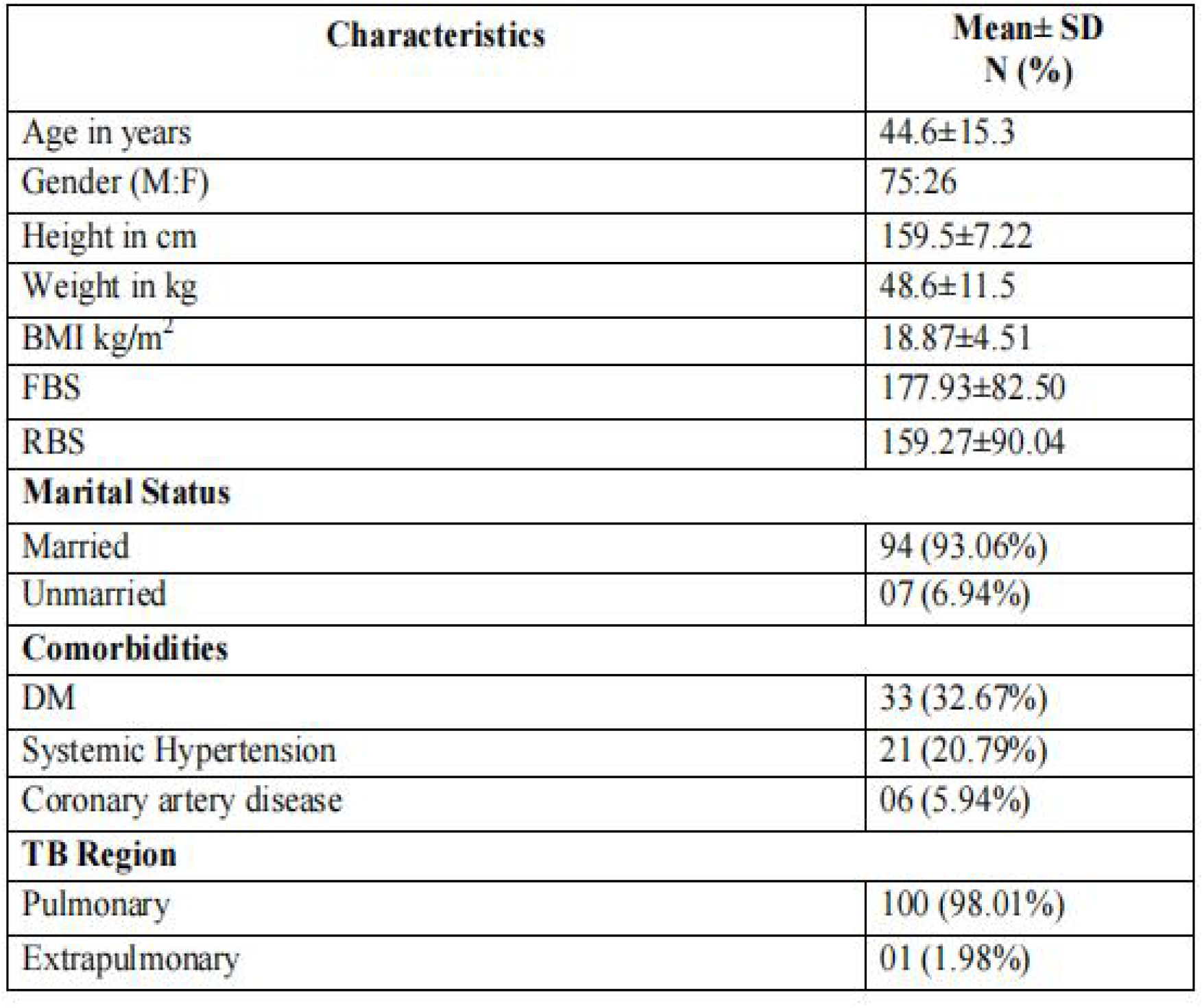
Baseline characteristics of the study population.

| Characteristics | Mean± SD<br>N (%) |
| --- | --- |
| Age in years | 44.6±15.3 |
| Gender (M:F) | 75:26 |
| Height in cm | 159.5±7.22 |
| Weight in kg | 48.6±11.5 |
| BMI kg/m<sup>2</sup> | 18.87±4.51 |
| FBS | 177.93±82.50 |
| RBS | 159.27±90.04 |
| **Marital Status** |  |
| Married | 94 (93.06%) |
| Unmarried | 07 (6.94%) |
| **Comorbidities** |  |
| DM | 33 (32.67%) |
| Systemic Hypertension | 21 (20.79%) |
| Coronary artery disease | 06 (5.94%) |
| **TB Region** |  |
| Pulmonary | 100 (98.01%) |
| Extrapulmonary | 01 (1.98%) |

#### Age

The study participants mean age was 44.6±15.3 years and ranged from 14 to 87 years. The predominant age group was between 40 to 50 years (Fig.1A.a).

**Figure 1A.**
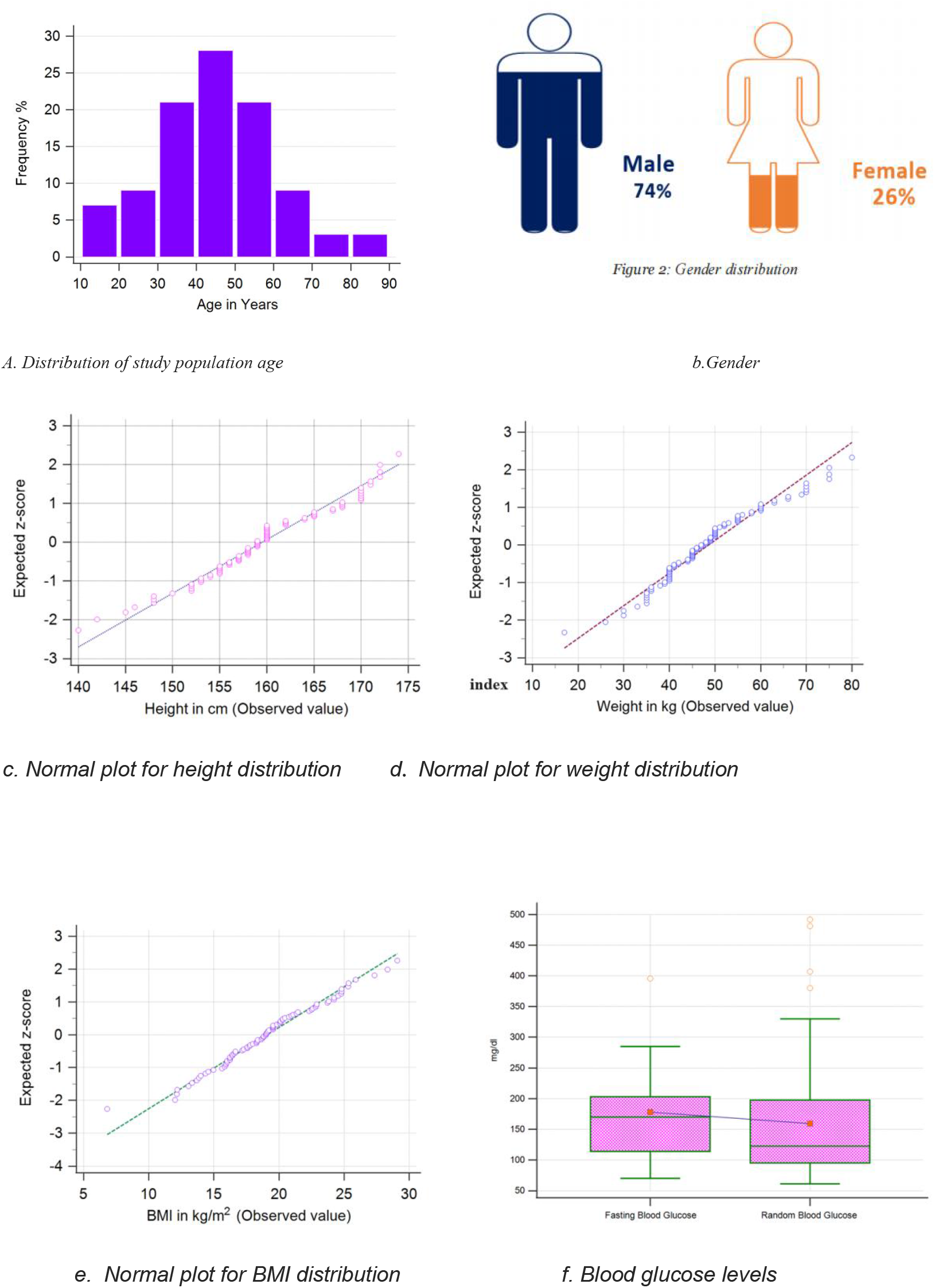
Demographics and clinical characteristics based on age, gender,height, weight, BMI and Glucose levels

#### Gender

The predominant gender was male (74%) and followed by the female (26%) observed in the study (Fig.1A b)

#### Height

The average height of the study participants was 159.5±7.22 cm and ranged from 140 to 174 cm (Fig1A c)

#### Weight

The weight of the study population was ranged from 17 to 75 kg with average of 48.6±11.5 kg. It showed that majority of the participants were lean body weight (Fig.1A d).

#### Body mass index (BMI)

The average BMI was 18.87±4.51 kg/m2 which showed a normal range. However, around 28 % of patients who had below normal BMI were observed in the study (Fig1A e).

#### Blood glucose

The blood glucose levels (Fig.1A f) were determined and found that the fasting blood glucose levels were higher (177.93±82.50 mg/dl) than random blood glucose levels (159.27±90.04 mg/dl)

#### Marital status

In the present study, the majority of the study population was married (93%) followed by 4% and 3 % of patients who were single and unmarried respectively (Fig1B a)

**Figure 1B.**
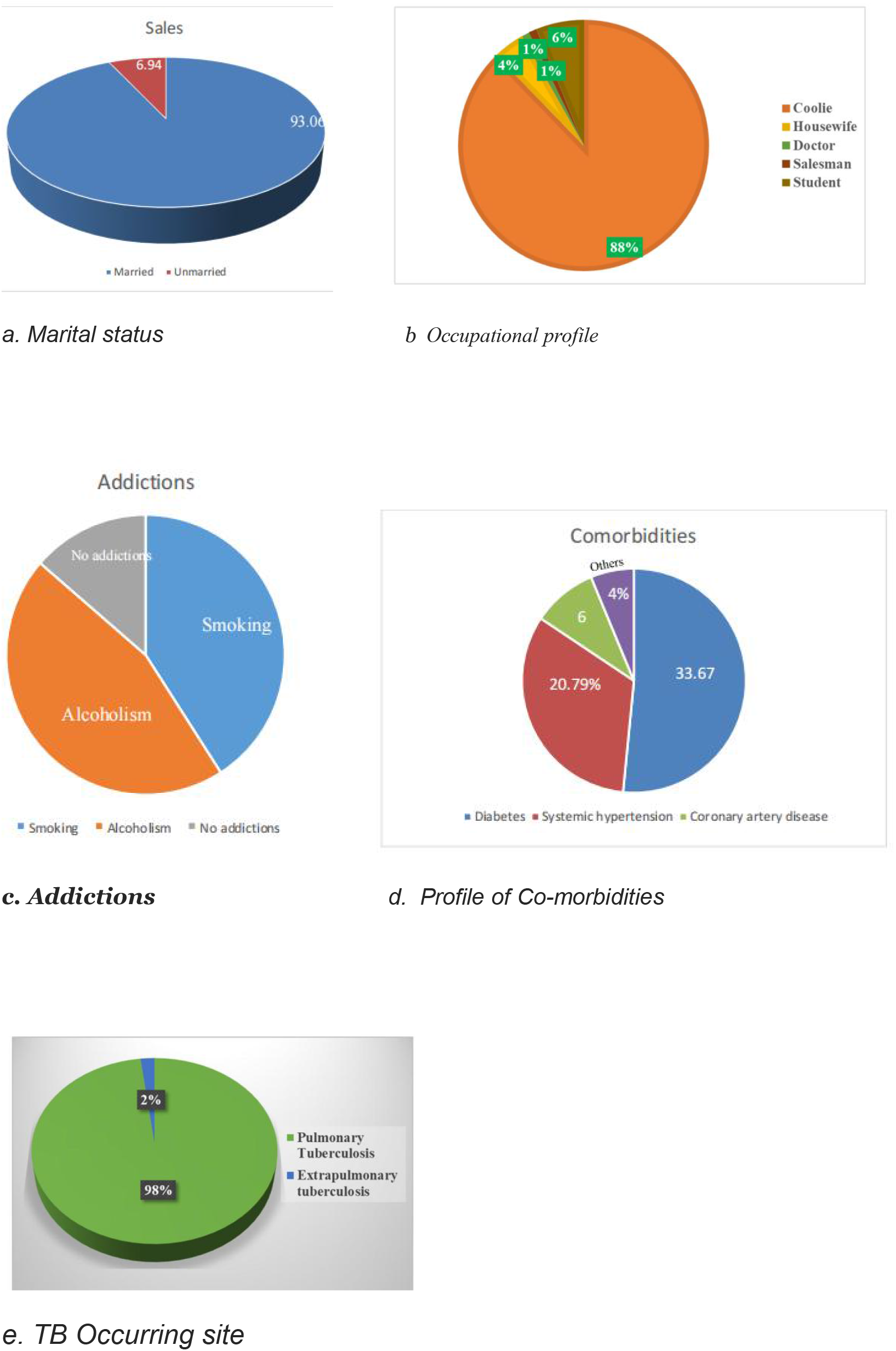
Demographics and clinical characteristics based on mariatal status, Occupational profile, addictions, profile of Co-morbidities and TB occuring site

#### Occupation

The predominant study population belongs to coolie (88%) followed by students (6%) and house-wife (4%). The remaining doctor and salesman were 1% each (Fig1B b)

#### Addictions

Smoking was found to be associated in 45% of the patients. Alcoholism was found in 50 % patients. No addictions was found in 15% patients. (Fig1B c)

#### Co-morbidities

*Diabetes mellitus* was the only co-morbidity observed in this study which was 33.67%, systemic hypertension 20.97%, coronary artery disease 6% and others 4% of the study population did not have any co-morbidities (Fig.1B d)

#### TB Site

In the study population, the majority of the patients had pulmonary tuberculosis (98.01%) and 1.98% of the population had extrapulmonary tuberculosis (Fig.1B e)

### Clinical Characteristics of the study population

#### History of anti-tuberculosis treatment

The history of previous ATT was observed in 43 patients (42.6%). Among 43 patients, sputum positive at diagnosis and retreatment was found in 31 patients (72.1%). Besides, 58 patients (57.4%) did not have any previous history of ATT.

#### Gene responsible

The gene responsible for INH monoresistant TB were KAT G (67.3%) and INH A (32.7%) of patients (Fig.2 a)

**Figure 2.**
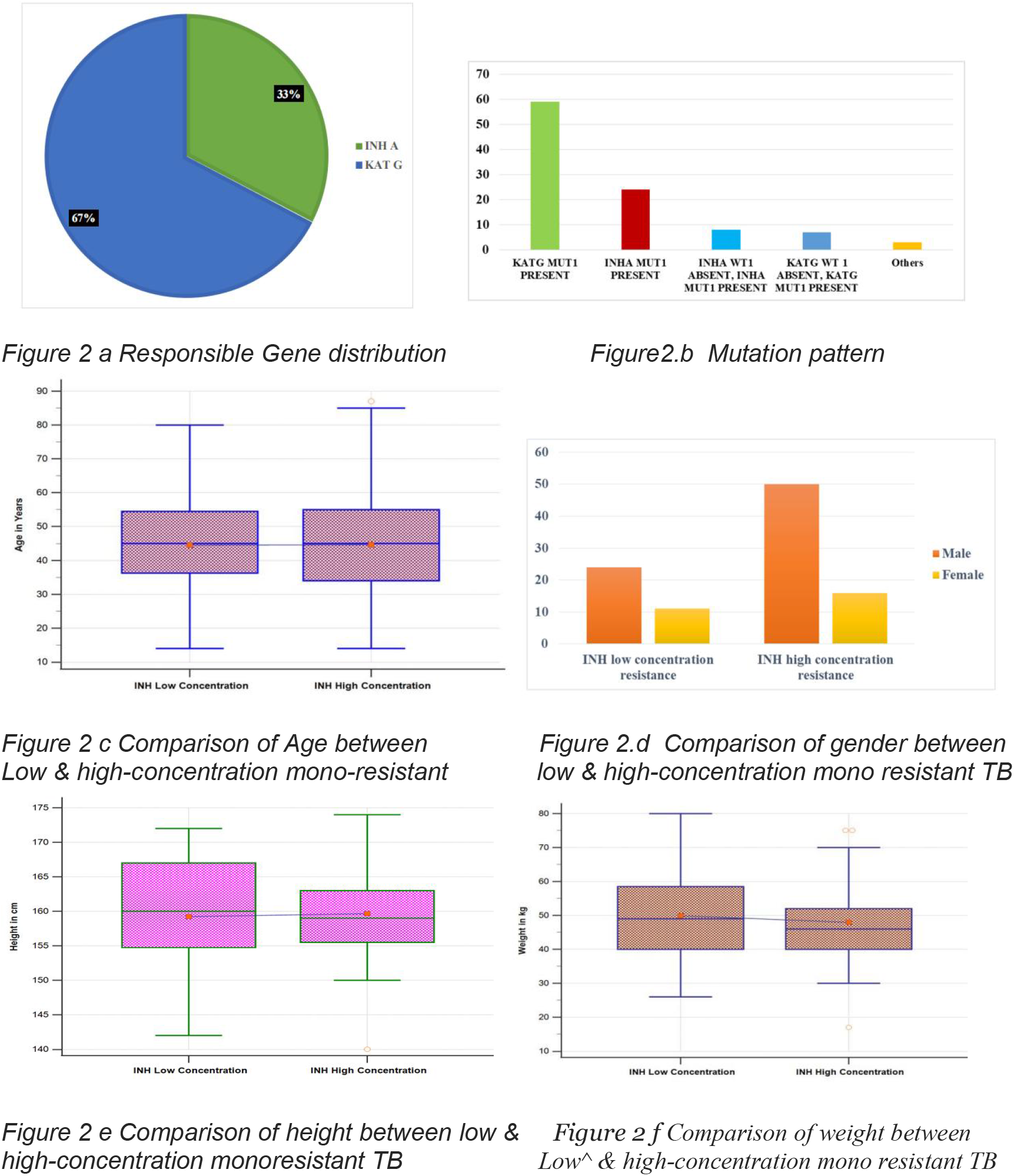
Comparision of age, gender, height and weight between low & high concentration monoresistant TB.

#### Mutation Pattern

The katG Mut1 was a prime mutation observed in the present study population (58.41%). Besides, inhA Mut1 was found in 23.76%, inhA Mut1 present &WT1absent in 7.9%, katG Mut1 present & WT1 absent in 6.9%, and other mutations were observed in 2.97% (Fig.2 b)

### Comparison of clinico-microbiological profile in INH low and high *concentration resistance*

The clinico-microbiological parameters were compared with the low and high levels of INH mono-resistance listed in table 2. The mean age of the patients with low- and high-concentration mono-resistant TB were 44.54±14.68 and 44.59±15.80 years respectively. It showed statistically not significant (P=0.9881) (Fig.2 c). Male gender was 68.6% and 75.75% in low- and high-concentration mono resistant TB respectively. The male gender was significantly higher in high concentration mono-resistant TB (p=0.0025) (Fig.15). Besides, the female gender was not significantly differed between the two groups of patients (p=0.3359). The average height of low and high concentration mono-resistant TB was 159.20±8.51and 159.68±6.53 respectively. Between both groups, the mean height was almost the same and was not statistically significant (p= 0.7772) (Fig.2 d)

**Table 2:** Demographic and clinical characteristics of the patients.

| Parameters | INH low<br>concentration<br>resistance<br>N=35 | INH high<br>concentration<br>resistance<br>N=66 | p Value |
| --- | --- | --- | --- |
| Age | 44.54±14.68 | 44.59±15.80 | 0.9881 |
| Male | 24 (68.6%) | 50 (75.75 %) | 0.0025 |
| Female | 11 (31.4 %) | 16 (24.24%) | 0.3359 |
| Height in cm | 159.20±8.51 | 159.68±6.53 | 0.7772 |
| Weight in kg | 49.93±12.04 | 47.92±11.25 | 0.4142 |
| BMI kg/m<sup>2</sup> | 18.82±5.42 | 18.9±4.02 | 0.9407 |
| H/O ATT | 15 | 28 | 0.0474 |
| Alcoholism | 6 | 18 | 0.0484 |
| Smoking | 4 | 10 | 0.0442 |
| Smoking and alcohol | 14 | 35 | 0.0213 |
| Pulmonary tuberculosis | 30 | 66 | 0.0002 |

The mean weight of low and high-concentration mono-resistant TB did not show any statistically significant difference (p=0.4142) (Fig.2 e). The mean body mass index (BMI) in low concentration mono-resistant TB was 18.82±5.42 and in high concentration mono-resistant TB was 18.9±4.02. The BMI in both groups has shown almost similar which was statistically no significant difference (p=0.9407) (Fig.3 a).

**Figure 3.**
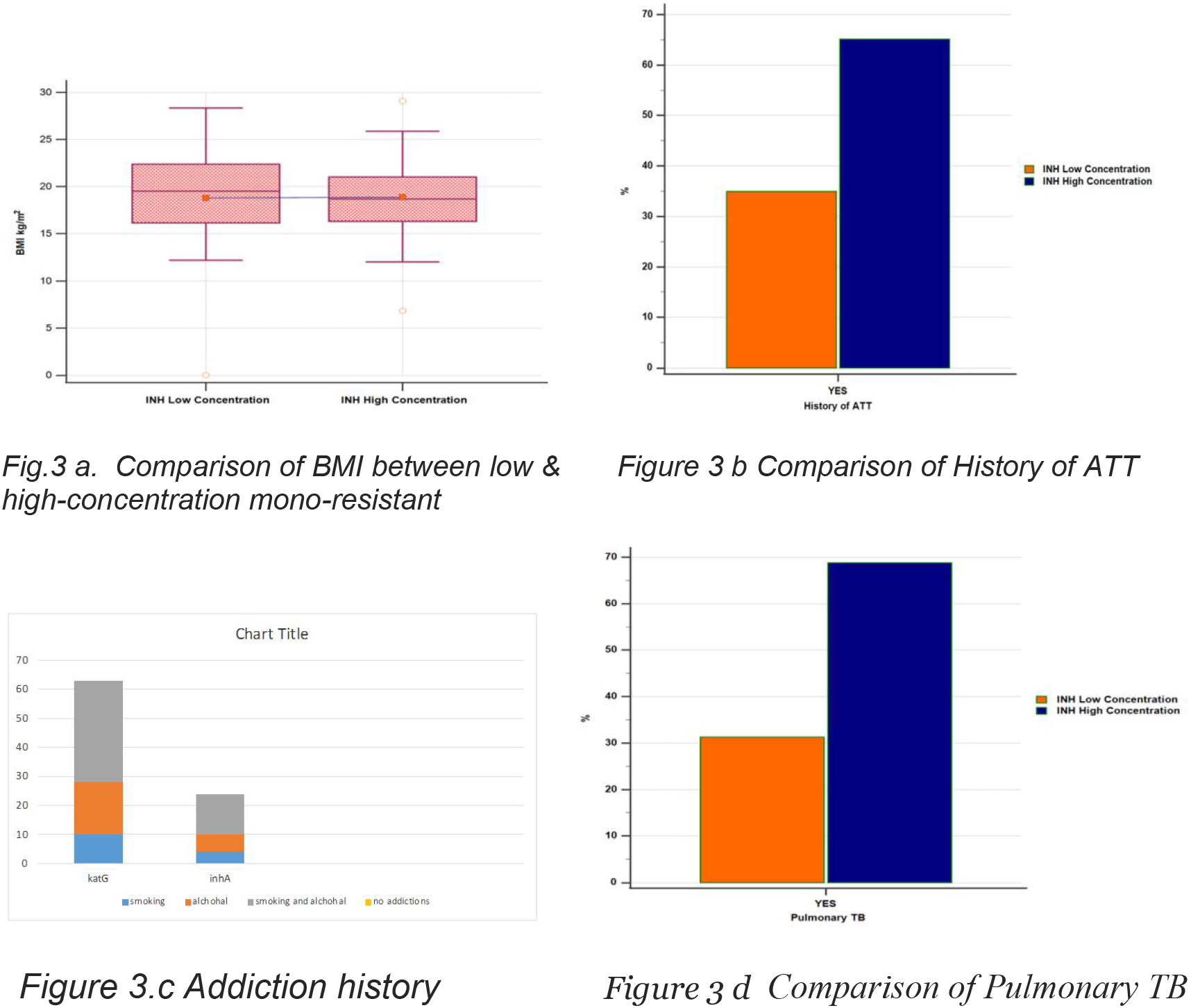
Comparison of BMI, ATT, Adiction and pulmanary TB.

The previous history of ATT was higher in high concentration resistance than in low concentration which was statistically significant (p=0.0474) (Fig.3 b). Smoking and alcoholism as addiction history was predominant and significantly higher in high concentrations than in low concentrations (p=0.0373) (Fig.3 c). The occurrence of TB in the pulmonary region was predominant and significantly higher in high concentrations than in low concentrations (p=0.0002) (Fig.3 d).

### Pattern of Gene mutation

The most common mutation in INH-resistant strains was in the ***kat***G gene (64.35%) followed by ***inh***A gene (34.65%). In 1 (1%) patient, both ***inh***A and ***kat***G gene mutations were observed (Table 3 & Fig.4). Comparing the pattern of gene mutations in INH monoresistant strains revealed that of 35 isolates with only ***inh***A gene mutation, WT1 pattern was absent and MUT1 present in 8 (22.9%) strains, WT2 pattern was absent and MUT1 present in 3 (8.6%), while MUT1 pattern was present in 24 (68.6 %) strains. ***Kat***G gene mutation patterns were observed in 66 isolates with different sequences. WT1 was absent and MUT1 was present in 6 (9.1%), inhAWT2 was absent and inhA MUT1 was present in 1 (1.5 %), MUT1 was seen in 58 (87.9%) patients (Table 4).

**Table 3:** Pattern of gene mutations in INH-resistant TB strains.

| Gene | N | % |
| --- | --- | --- |
| inhA | 35 | 34.65 |
| katG | 65 | 64.35 |
| Both | 01 | 1 |

**Table 4:** Analysis of patterns of gene mutations at various loci of InhA and katG in Isoniazid Mono-resistant Tuberculosis Patients.

| Gene | INH low concentration resistance<br>N=35 |  |  |  | INH high concentration resistance<br>N=66 |  |  |  |
| --- | --- | --- | --- | --- | --- | --- | --- | --- |
|  | Locus |  |  |  |  |  |  |  |
|  | WT1(-) | WT2 (-) | MUT1 (+) | MUT2 (+) | WT1(-) | WT2 (-) | MUT1 (+) | MUT2 (+) |
| inhA | 8 | 3 | 35 | - |  | 1 | 1 |  |
| KatG | - | - | - | - | 6 |  | 60 |  |

**Figure 4A.**
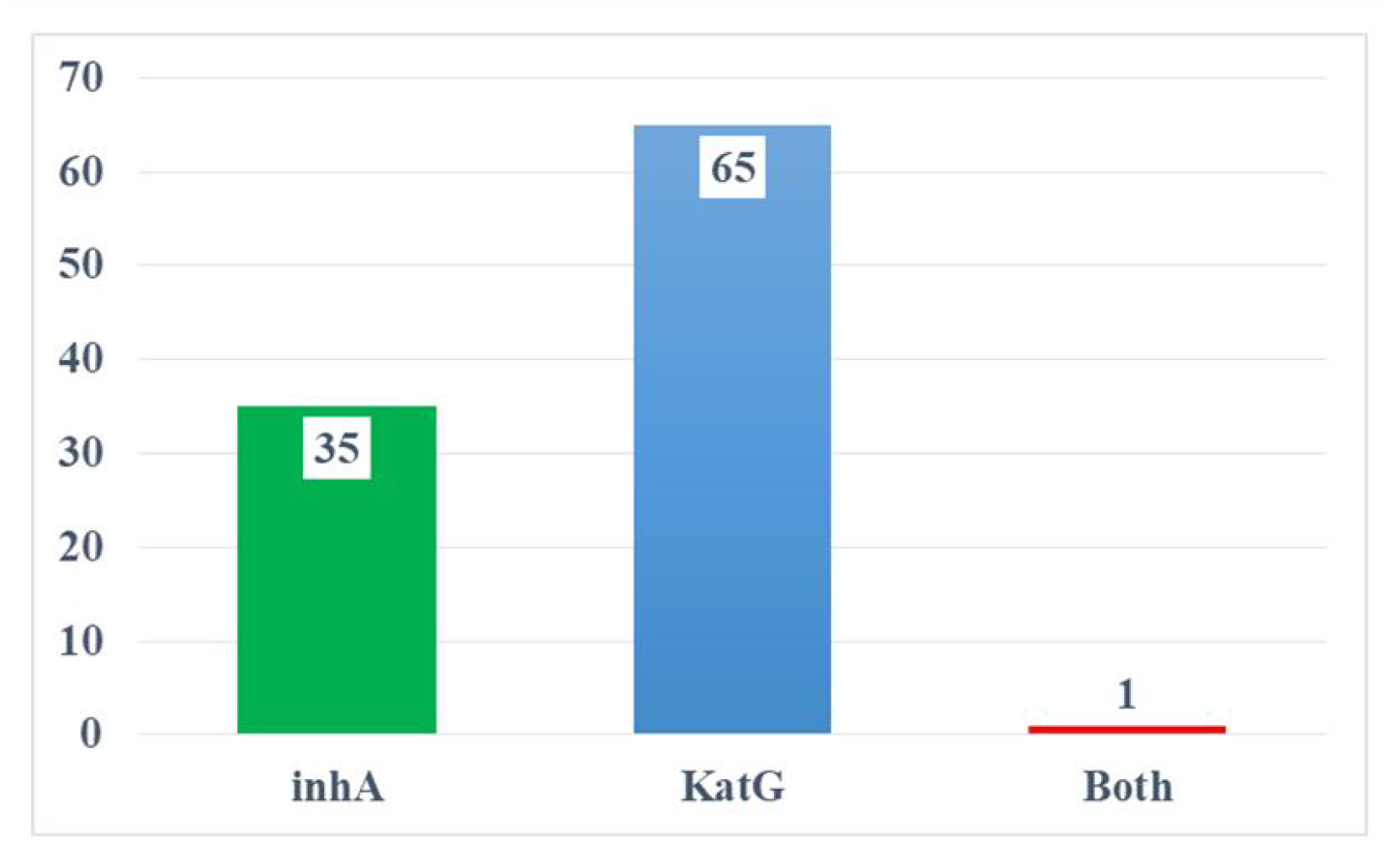
Pattern of gene mutations.

**Figure 4B.**
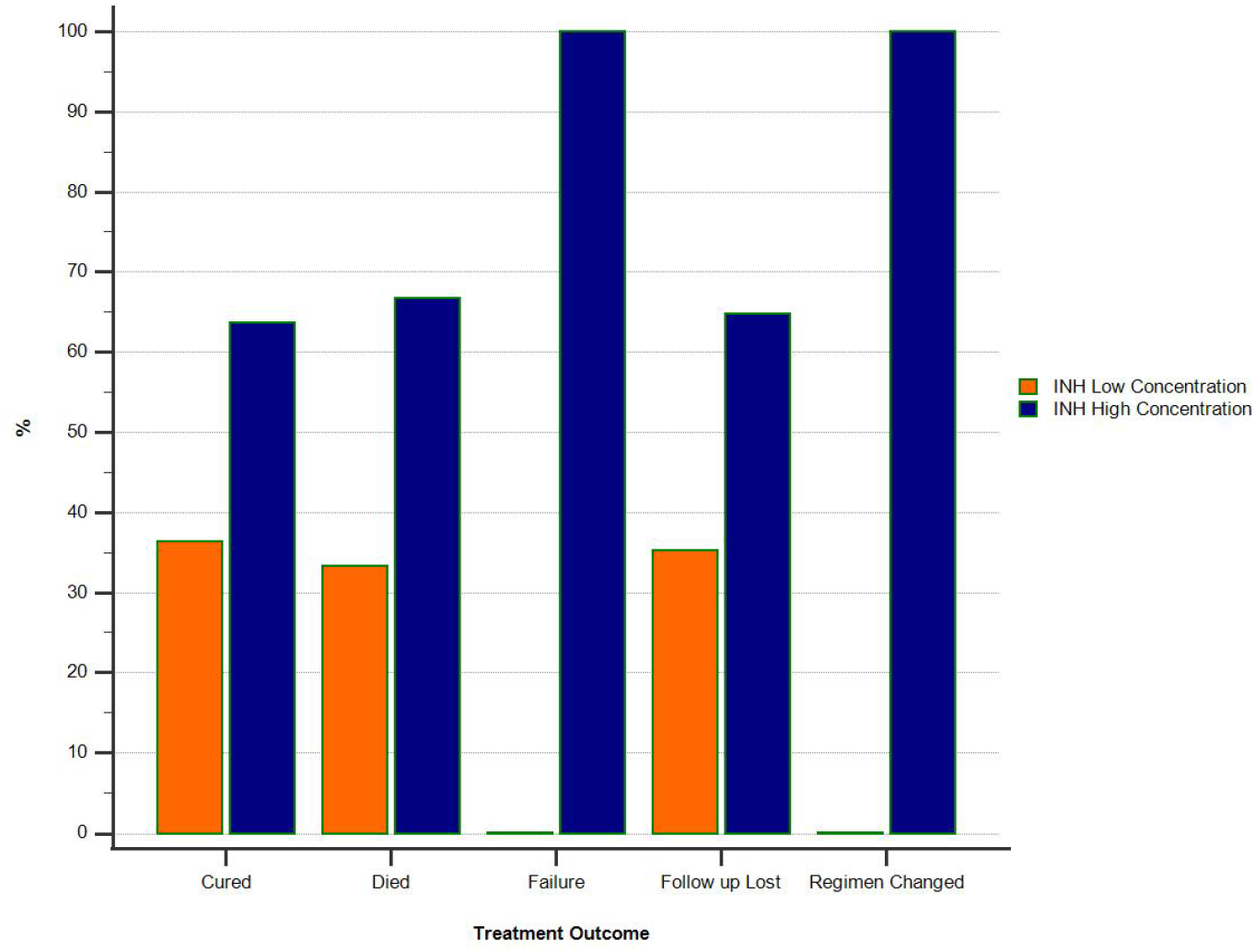
Treatment outcome between two groups.

### Treatment outcome

Out of 101 INH monoresistant tuberculosis patients, 35 had low-dose concentration resistance (inhA), of which 28 patients were cured. 66 patients had high dose resistance (katG), of which 49 patients were cured. Cure rate was high in *INH high concentration resistance* patients which were statistically significant (p=0.0167) (Fig.5)

**Figure 5A.**
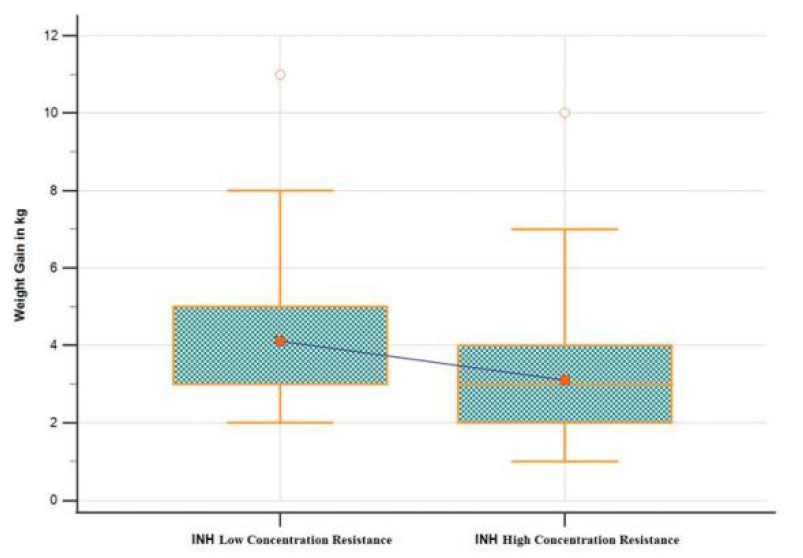
Comparison of weight gain.

**Figure 5.B.**
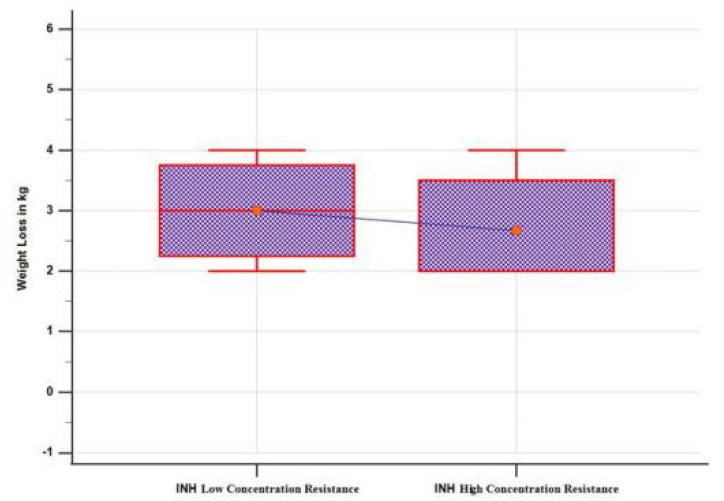
Comparison of weight loss.

### Treatment impact on weight

The treatment impact on patients’ weight was determined and found that weight gain was a major outcome in both low and high concentrations (Table 6). However, the weight gain was not statistically significant (p=0.0622) (Fig.6 A). Similarly, the weight loss was also statistically not significant (p=0.7247) (Fig.6 B). Besides, there were no changes in weight after treatment was observed in some patients but it was not significantly differed.

**Table 5:**
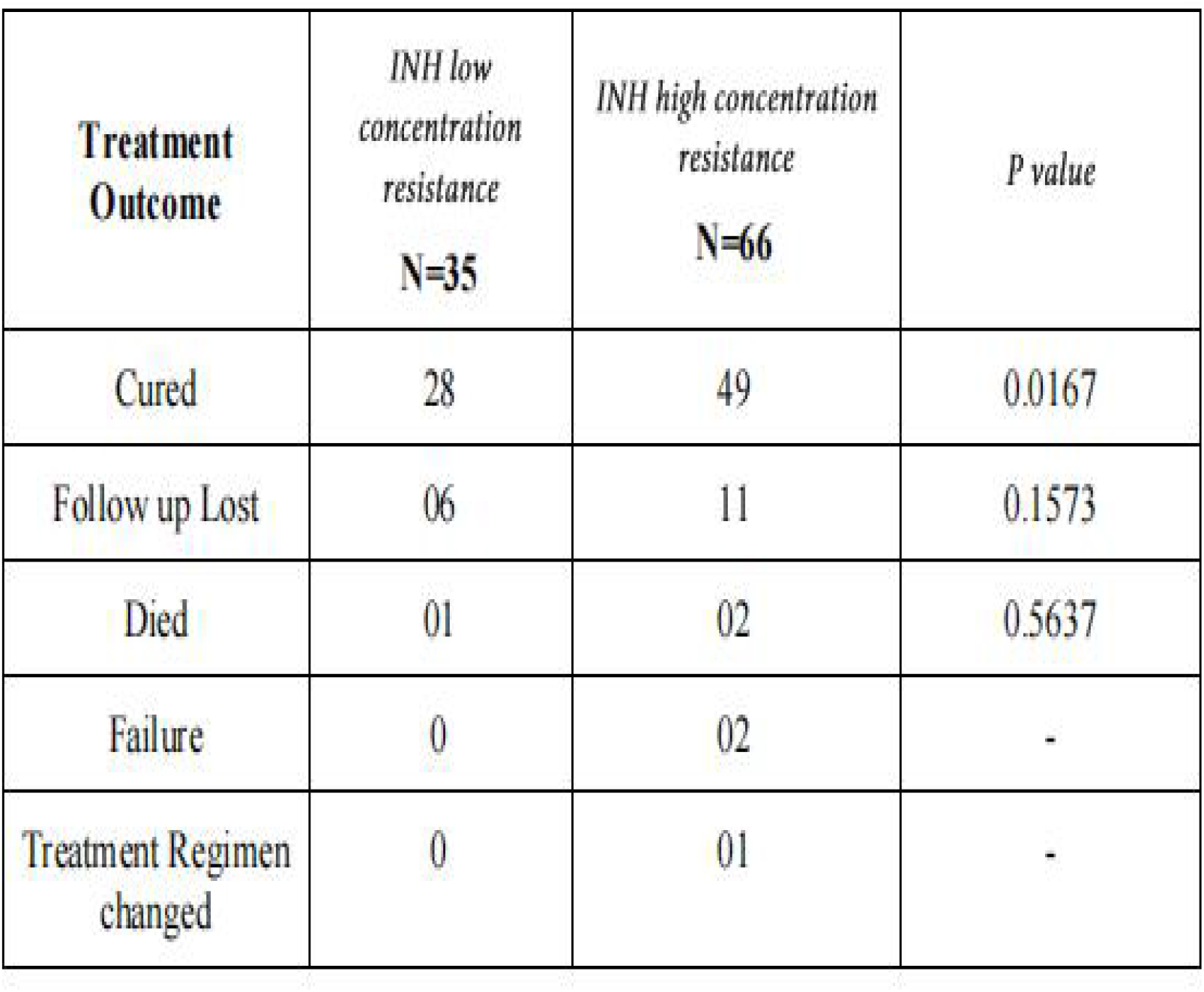
Treatment outcome between two groups.

**Table 6:** Impact of Treatment on weight.

| **Weight Status** | *INH low concentration resistance*<br>**N=35** | *Mean±SD* | *INH high concentration resistance*<br>**N=66** | *Mean±SD* | **p Value** |
| --- | --- | --- | --- | --- | --- |
| Weight Gain | 19 (54.3%) | (4.10±2.23 kg) | 41(62.12%) | 3.09±1.74 | 0.0622 |
| Weight Lost | 3 (8.6%) | (3±1 kg) | 3(4.54%) | 2.7±1.1 | 0.7247 |
| No change | 7 (20%) | - | 8 (12.12%) | - |  |

**Figure 6:**
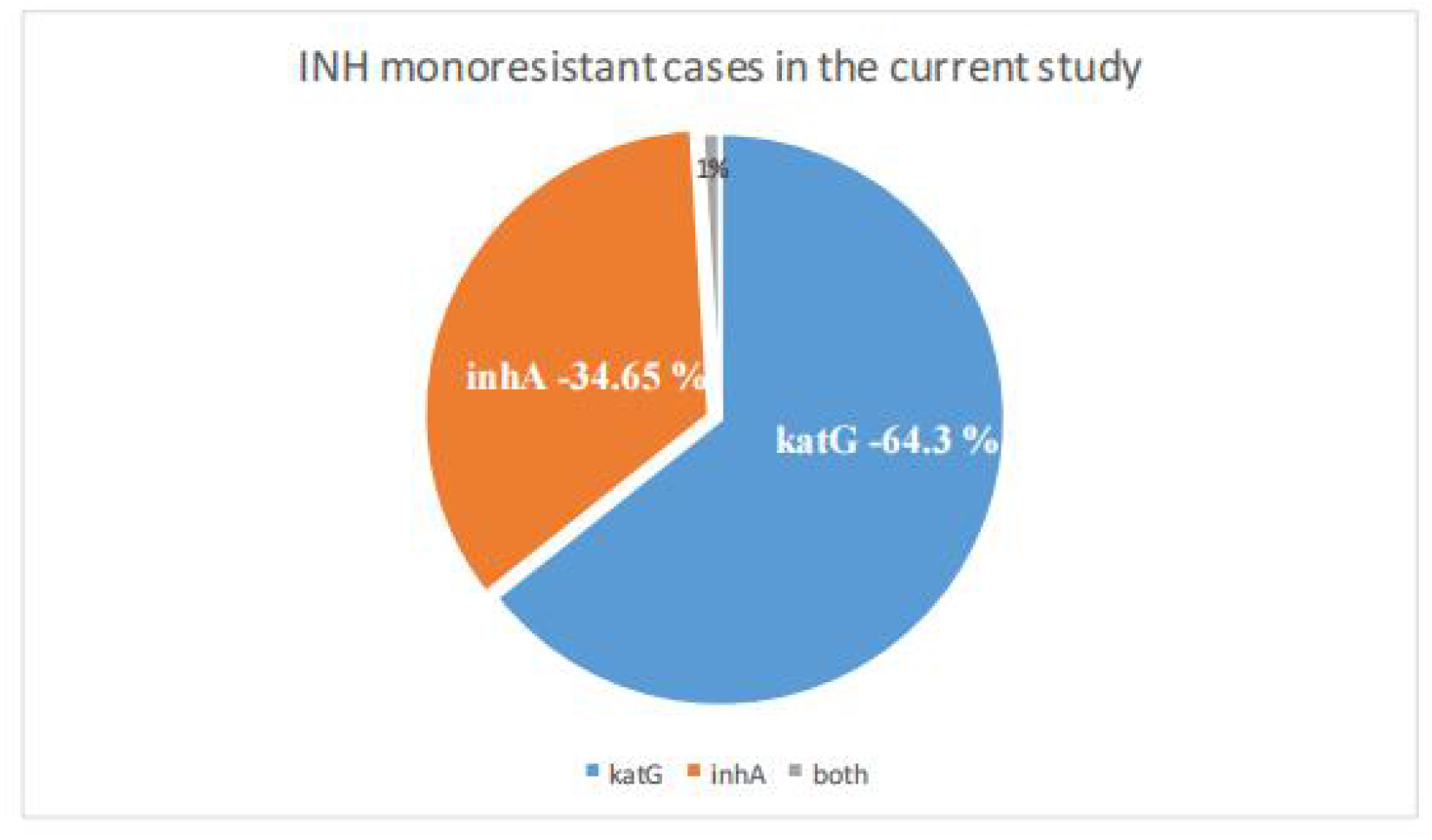
Current study showing gene association with INH monoresistance.

## DISCUSSION

As a result of its effective early bactericidal activity against rapidly proliferating cells, isoniazid is a crucial first-line anti-TB medication [22]. Isoniazid helps to prevent the selection and formation of a drug-resistant TB population when used in combination with other anti-TB medications. But among all first-line medications, isoniazid resistance is the most prevalent, with an estimated frequency of 13% globally, including new and retreatment cases [23]. It is still unknown what the best treatment plan is for isoniazid mono-resistant TB. According to several recent studies, [24,25] individuals with isoniazid mono-resistant TB fared worse than those with drug-susceptible TB, despite studies from the 1970s and 1980s reporting a low rate of treatment failure for those receiving four or five medications over a six month period.

According to the results of the current investigation, *Diabetes mellitus* affected 33% of the patients. Recedntly, West Bengal study [26] found that 43% of TB patients also had diabetes. A study conducted in Indonesia, Peru, South Africa and Romania with a high TB burden found that 12.5% of TB patients had diabetes [27]. The WHO strategy of routine bidirectional symptom-based TB screening in known DM patients in high TB burden countries has been endorsed by Alisjahbana et al.[28]. In our current study Diabetes patients having 54 history of previous tuberculosis and ATT intake are having high dose isoniazid monoresistance (katG). The people who had arrived for the presumed TB test were overwhelmingly male. Male patients made up 74% of those with MTB positivity. Our results are in line with those of recent studies in India that found that 76%, 75%, 70.5%, 71%, and 72% of TB patients were males, as reported [29–32]. According to the WHO study from 2021, adult males, who made up 56% of all TB cases in 2019, have the highest prevalence of the disease [6]. More than one gene or gene complex, including the katG, inhA, and kasA genes as well as the intergenic region of the oxyR ahpC complex, may be altered in the genetic foundation of INH resistance [33]. Two genes, katG and inhA, have probes that can be used to identify INH resistance in Genotype MTBDR +. According to Van Rie et al.,[34] the katG mutation was less common (37.6%). Numerous studies have demonstrated a correlation between high and low levels of INH resistance and the codon 315 of the katG gene (50%–90%) and regulatory region of the inhA gene (20%– 35%), respectively[35]. In our study we found that the katG gene (67% of patients had this mutation), followed by the inhA gene (33% of patients). Yao et al. (2010) obtained results that were almost identical. A 50 INH monoresistant patients were examined, and it was found that 41 (82%) had KatG mutations and 9 (18%) had inhA mutations [36]. In addition, according to Huyen et al., 28.2% and 75.3%, respectively, of the population, contained mutations in the inhA promoter region [37]. Similar findings were made by Kigozi et al. (2018) who found that 6% of samples had inhA mutations and 80% had katG mutations [38].

In addition, Tavakkoli and Nazemi (2018) reported that inhA genes and katG genes, respectively, were in charge of INH resistance in 17.24% and 82.76% of the strains [39]. Niehaus et al. discovered that 33.1% of 924 isolates, or 30.3% of those with MDR TB, 47.2% of those with pre-XDR-TB, and 82.8% of those with XDR-TB, carried an inhA mutation with or without a katG mutation [40]. Alagappan et al. (2018) looked into the promoter region of the inhA gene and codon katG for INH resistance mutations in M. tuberculosis. Of the 15,438 INH resistant bacteria, 1,821 (11.8%) showed detectable mutations, with 71.0% occurring in katG315 and 29.0% in the inhA promoter region[41]. Isakova et al. (2018) found that the katG gene was mutated in 91.2% of strains, the inhA gene was mutated in 7% of specimens, and the ahp C gene was mutated in 2 more specimens (1.8% of the total) [42]. Jagielski et al. (2015) also noted that katG mutations were present in 85.2% of MDR patients. Approximately 3.7% of the 54 patients had only one inhA mutation, and the remaining patients, or 11.1%, had both mutations [43]. A comparative outcomes.are shown in Table 7.

Clinicians should be aware of mutations in the katG or inhA promoter region.[44] and high level of resistance to INH is indicated by the existence of mutations in katG alone or in conjunction with inhA. For these patients, adding even high doses of INH is unlikely to improve the efficacy of a regimen. On the other hand, a mutation restricted to inhA is typically associated with a low level of INH resistance, and these people are likely to benefit from large doses of INH (10–15 mg/kg/day) [45]. Results of our study showed mean age of incidence with low- and high concentration mono-resistant TB were 44.54±14.68 and 44.59±15.80 years respectively. It showed statistically not significant (p=0.9881). Besides, the male gender was 68.6% and 75.75% in low-and high-concentration mono-resistant TB respectively. The average height of the study participants was 159.5±7.22 cm and ranged from 140 to 174 cm. The weight of the study population was ranged from 17 to 75 kg with average of 48.6±11.5 kg. It showed that majority of the patients in the study had low BMI. The predominant study population belongs to coolie (88%) followed by students (6%) and housewife (4%). The remaining doctor and salesman were 1% each. *Diabetes mellitus* was the most common co-morbidity observed in this study which was 33% in the population. Smoking and alcohol association was associated with patients diagnosed with high dose INH monoresistant tuberculosis (53.03%) of which significant of the patients were associated with retreatment (68.57%). In the study population, most of the patients had pulmonary tuberculosis (98.01%) and 1.98% of the population had extrapulmonary tuberculosis (pleural effusion). The history of previous ATT was observed in 43 patients (42.6%). Among 43 patients, sputum positive at diagnosis and retreatment was found in 31 patients (72.1%). Besides, 58 patients (57.4%) did not have any previous history of ATT. INH high dose monoresistance was seen in 66 patients and low dose mono resistance was 58 seen in 35 patients. The predominant gene responsible for TB was katG (64.3%) and inhA gene was observed in 34.65% of patients. The katG Mut1 was a prime mutation observed in the present study population (58.41%). Besides, inhA Mut1 was found in 23.76%, inhA Mut1 present &WT1 absent in 7.9%, katG Mut1 present & WT1 absent in 6.9%, and other mutations were observed in 2.97%. Treatment outcomes showed 77 patients were cured (74.75%), 17 patients were lost to follow up (16.5%), 3 patients died during the course of treatment (2.91%), treatment failure was seen in 2 patients (1.94%), treatment regimen changed in 1 patient (0.97%). Study showed positive association with males (74%), occupation as coolie (88%), diabetes as comorbidity (33%), pulmonary tuberculosis as the TB site (98.01%), history of previous ATT intake in 43 patients (42.6%), katG mutation (67.3%), katG Mut 1 was the prime mutation (58.4%). Study also showed that male patients with diabetes has comorbidity, with history of ATT intake along with sputum positive at diagnosis and retreatment (72.1%) were associated with INH high dose monoresistance (katG), having katG Mut 1 as prime mutation. The importance of treating physicians being aware of these gene alterations as well as the usefulness of laboratories reporting these mutations cannot be overstated.

## CONCLUSION

Patients with both smoking and alcohol addictions were with INH high dose monoresistance (katG), of which katG Mut 1 prime mutation was predominant (53.03%). A 32 patients with history of previous tuberculosis were diagnosed to have recurrent tuberculosis with high dose INH monoresistance out of which 27 had diabetes, smoking and alcohol association (84.3%). Current study showed that there is high prevalence of INH monoresistance in patients with previous H/O ATT intake, alcohol, smoking and diabetes. Results of the study showed that there is a significant association between males, diabetes, smoking and alcohol addictions were associated with high risk of developing high dose INH monoresistance (katG)In Pondicherry, due to effective screening, implementation of NTEP program and there has been a rise in incidence of INH monoresistance tuberculosis in past few years raising a concern for increase in risk of treatment failure and multi drug resistant tuberculosis. Present study conclude that high prevalence of addictions, prevailing low socioeconomic class had influence in treatment outcomes, incidence of recurrent cases and high risk for development of INH monoresistance tuberculosis in Puducherry. Patients who are microbiologically confirmed pulmonary tuberculosis and diabetes with rifampicin sensitive status needs to be checked for LPA for isoniazid sensitivity status to prevent treatment failure and relapse. It is crucial to understand the gene pattern in each of these patients since these mutations are closely associated to high or low-degree resistance to INH.

## Data Availability

the data used for analysis is from drug TB registry from Intemediate reference laboratory, Puducherry

## Acknowledgments

We would like to acknowledge Dr. Muthuraj R, chief microbiologist, Intermediate reference laboratory, Puducherry, for his support in providing laboratory data and Dr Ezhumali for his statistical assistance.

## Author Contributions

**Conceptualization:** Pajanivel R, Venkata Prasanna M

**Data curation:** Venkata Prasanna M

**Formal analysis:** Pajanivel R, Venkata Prasanna M, Muthuraj R, Vimal Raj R

**Methodology:** Pajanivel R, Venkata Prasanna M, Muthuraj R

**Software:** Venkata Prasanna M

**Supervision:** Pajanivel R, Muthuraj R, Govindarajan S, Vimal Raj R

**Visualization:**Venkata Prasanna M, Pajanivel R, Muthuraj R, Govindarajan S, Vimal Raj R

**Writing** –Venkata Prasanna M, Pajanivel R

**Original draft:** Venkata Prasanna M, Pajanivel R, Muthuraj R, Vimal Raj R

**Writing – review & editing:** Venkata Prasanna M, Pajanivel R, Muthuraj R, Govindarajan S, Vimal Raj R

